# Protocol of a randomized controlled trial evaluating program facilitating empathetic communication for initiating advanced care planning discussions between patients with advanced cancer and healthcare providers (J-SUPPORT 2104)

**DOI:** 10.1101/2022.11.15.22281288

**Authors:** Kyoko Obama, Maiko Fujimori, Masako Okamura, Midori Kadowaki, Taro Ueno, Narikazu Boku, Masanori Mori, Tatsuo Akechi, Takuhiro Yamaguchi, Shunsuke Oyamada, Ayumi Okizaki, Tempei Miyaji, Naomi Sakurai, Yosuke Uchitomi

**Affiliations:** Institute for Cancer Control, National Cancer Center, Tokyo, Japan; SUSMED, Inc, Tokyo, Japan; Department of Oncology and General Medicine, IMSUT Hospital, The Institute of Medical Science, The University of Tokyo, Tokyo, Japan; Division of Palliative and Supportive Care, Seirei Mikatahara Hospital, Hamamatsu, Japan; Department of Psychiatry and Cognitive-Behavioral Medicine, Nagoya City University Graduate School of Medical Sciences, Nagoya, Japan; Division of Biostatistics, Tohoku University Graduate School of Medicine, Sendai, Japan; Department of Biostatistics, JORTC Data Center, Tokyo, Japan; Cancer Survivors Recruiting Project, General Incorporated Association, Tokyo, Japan

**Keywords:** Advance care planning, Empathetic communication, Question prompt list, Advanced cancer, Randomized controlled trial

## Abstract

**Introduction:** In patients with incurable advanced cancer, preferences about treatment and how to spend their final days are not adequately discussed. This process of discussion is called “advanced care planning” (ACP), and timely intervention is recommended. As the communication attitude of healthcare providers is a critical factor in ACP facilitation, improving their communication attitudes may reduce patient distress, improve care satisfaction, and reduce unnecessarily aggressive treatment. Digital mobile devices are being developed for behavioral interventions due to their low space and time restrictions, and the ease of sharing information. The purpose of this study is to evaluate the effectiveness of a facilitation program utilizing a mobile app developed to improve communication between cancer patients and healthcare providers regarding ACP.

**Methods and analysis:** This study utilizes a parallel-group, evaluator-blind, randomized controlled trial design. We plan to recruit 264 adult patients with incurable advanced cancer. Intervention group participants will use a mobile app ACP program and undergo a 30-minute interview with a trained intervention provider to discuss with the oncologist at the next patient visit. Control group participants will continue their usual treatment. The primary outcome is the oncologist’s communication behavior score assessed using audio recordings of the consultation. The secondary outcomes include communication between patients and oncologists and the patients’ distress, quality of life, care goals and preferences, and medical care utilization. We will use a full analysis set with the registered participant population who received at least a part of the intervention.

**Ethics and dissemination:** The study protocol was reviewed and approved by the Scientific Advisory Board of the Japan Supportive, Palliative and Psychosocial Oncology Group (Registration No. 2104) and the Institutional Review Board of the National Cancer Center Hospital (registration No. 2020-500). The results of the RCT will be published in peer-reviewed scientific journals and presented at scientific meetings.

**Strengths and limitations of this study:**

- Randomized controlled trials using mobile apps for behavior change and psychological interventions are increasing, and this study is unique in its focus on facilitating communication about advanced care planning (ACP).
- The intervention will include mobile apps which can be used in environments the participants find relaxing and engaging. The benefit is particularly significant for patients with advanced cancer who need to express their values and what is crucial to them.
- There is currently no gold standard for evaluating ACP discussions between patients and healthcare providers. The methods facilitate ACP discussions in various aspects of this study to produce a variety of practical insights.
- The timing of introducing ACP discussions must be individualized to each patient, and it is anticipated that some participants may find this intervention burdensome. Therefore, more careful ACP referrals are needed, and qualitative exploration of study dropouts is needed.
- Multiple intervention components make it difficult to determine which is most effective. Individualized assessments of app usage, intervention adherence, and patient satisfaction could clarify the challenges and help determine next steps.

## INTRODUCTION

Cancer is a leading causes of death in developed countries, with an estimated 10 million cancer deaths worldwide in 2020, [1] accounting for a one-in-six risk of dying from cancer. However, healthcare providers do not adequately discuss treatment preferences with patients with incurable advanced cancer, nor how they may spend their final days. [2] Delayed discussions, such as after the patient’s condition deteriorates, are associated with unprofitable treatment and delayed coordination with community health services. [3] Communicating with patients with incurable advanced cancer is a considerably challenging tasks, especially regarding preferred end-of-life care appropriate to their treatment, although discussion helps patients and their families prepare for the end of life. Patients receiving communication intervention are more likely to have shared their end-of-life care preferences with healthcare providers, and consequently expressed higher satisfaction with them. [4] This discussion, called advanced care planning (ACP), is practiced based on clinical guidelines worldwide. [5–7] based on clinical guidelines. The National Comprehensive Cancer Network (NCCN) guidelines recommend beginning the ACP discussion when a patient’s estimated prognosis is one year or less. [8]

Since barriers to ACP include a lack of supportive and empathetic attitudes and inadequate information delivery by healthcare providers, [9] improving healthcare providers’ communication attitudes toward patients is essential for facilitating ACP. Likewise, ACP may improve communication regarding end-of-life care between cancer patients and healthcare providers [10–13] and make palliative care more accessible to patients. [14] Thus, ACP may reduce patients’ anxiety and depression, [15] increase satisfaction with care, [15] and reduce unnecessarily aggressive treatment. [16] The ACP intervention components include communication support using question prompt lists (QPL) for patients, [10 12 17] communication skill training (CST) for healthcare providers, [15 18] a combination of CST for healthcare providers and patients, [19] and step-by-step in-depth counseling to patients by trained facilitators. [14 20]

We previously developed a face-to-face behavioral intervention program using QPL and CST to improve the introduction of ACP discussion between healthcare providers and their cancer patients being given bad news. [21] Combining a 2.5 hr individualized communication skills training for healthcare providers with a 30 min coaching intervention for patients, we found statistically significant improvements in empathic communication and information sharing among healthcare providers. In addition, patients in the intervention group were more satisfied with the consultation. [21 22] However, from the perspective of implementing a communication intervention, face-to-face programs held in hospitals could create significant time and space burdens for both patients and healthcare providers.

To overcome these problems, we developed an ACP program mobile application (app). We revised the intervention program [21] to include an app with reference to previous QPL studies, [23–25] the goal concordant care framework, [26] the good death, [27 28] and digital health-based intervention. [29] Because of the advantages of digital health-based interventions, such as fewer space and time constraints and easier real-time information sharing compared to face-to-face interventions, more medical apps are being developed for behavioral interventions such as physical activity [27 28] and psychoeducation [30] among cancer patients. Intervention via apps can reduce the chance of patient contact, which is useful in the COVID-19 pandemic. The study’s purpose is to evaluate the effectiveness of a facilitation program utilizing a mobile app for improving communication between cancer patients and ACP healthcare providers regarding.

## METHODS AND ANALYSIS

### Study design

This study comprises a parallel-group, evaluator-blind, randomized controlled trial.

### Patient and public involvement

A cancer survivor from a patient advocacy group participated, giving suggestions regarding the study design and materials that were completed via a series of reviews. The study protocol has been reviewed by researchers, healthcare providers, patients, and the public through the Scientific Advisory Committee of the Japan Supportive, Palliative and Psychosocial Oncology Group (J-SUPPORT, the study ID: 2104). Five cancer patients who were attending a study field hospital volunteered to participate in the pretest; their comments were used to refine the study procedures.

### Study population

Participants are recruited from the Departments of Oncology, Hepatobiliary Medicine, Respiratory Medicine, and Gastroenterology at the National Cancer Center Hospital (Tokyo), Japan. Inclusion criteria are patients 20 years or older with incurable advanced cancer whose attending oncologist has indicated that they meet the Surprise Question [15 31] (answering “no” to the question “Would you be surprised if this patient die within a year?”); they must have an Eastern Cooperative Oncology Group performance status score 0–2, provide written consent to participate in the study, and be able to read, write, and understand Japanese. Exclusion criteria are patients who have been judged by the attending oncologist to have a serious cognitive decline, such as delirium or dementia, an estimated prognosis of fewer than 3 months, judged by an attending oncologist to be unsuitable for this study, participating in other psychological or communication support interventions at the time of enrollment.

### Enrollment and randomization

Participant management, including enrollment, randomization, and data collection via Electronic Patient-Reported Outcome (ePRO) and PRO, will be conducted online using the central registration system linked to the app developed in collaboration with SUSMED, Inc (Tokyo, Japan), a medical app developer. Research assistants explain the research to the candidates and obtain written consent. After obtaining baseline data, participants are randomly assigned using a minimizing method to either an intervention group or a usual care group in 1:1 with stratification factors of the clinical department (respiratory medicine, gastroenterology, hepatobiliary medicine, and oncology), gender (male and female), and age (at age 64 years or younger and 65 years or older). Allocation results are blinded to the primary outcome evaluators.

Detailed allocation procedures will not be shared with researchers at participating sites, data centers, or statistical analysts, and will be defined in an internal document at the site of the person responsible for allocation. Participants will install the app on their mobile devices upon enrollment. Participants allocated to the control group will use an app that contains only ePRO, while the intervention group will use an app that contains the intervention program in addition to ePRO. If the app cannot be installed on the participant’s mobile device for some reason, an iPad with the app installed will be available for loan.

### Procedures

Five visits are planned: baseline evaluation (T0), an outpatient visit at least one week later (T1), and follow-up surveys at 1 week (T2), 12 weeks (T3), and 24 weeks (T4) after the T1 visit, as shown in Figure 1. The purpose of each visit was mainly to evaluate how the intervention program impacts communication between participants and their oncologists during the consultation at T1, the psychological burden of the participants around 2 weeks after the consultation at T2, and the patients’ preferred end-of-life care settings and care preferences and their actual healthcare utilization at T3 and T4. Intervention group participants receive interventions before T1. Usual care (control) group participants receive care as usual. The schedule for outcome measurement is shown in Table 1. At the T1 visit, the consultation is audio recorded. The research assistant reminds and asks participants to respond to ePRO according to the response schedule.

**Figure 1.**
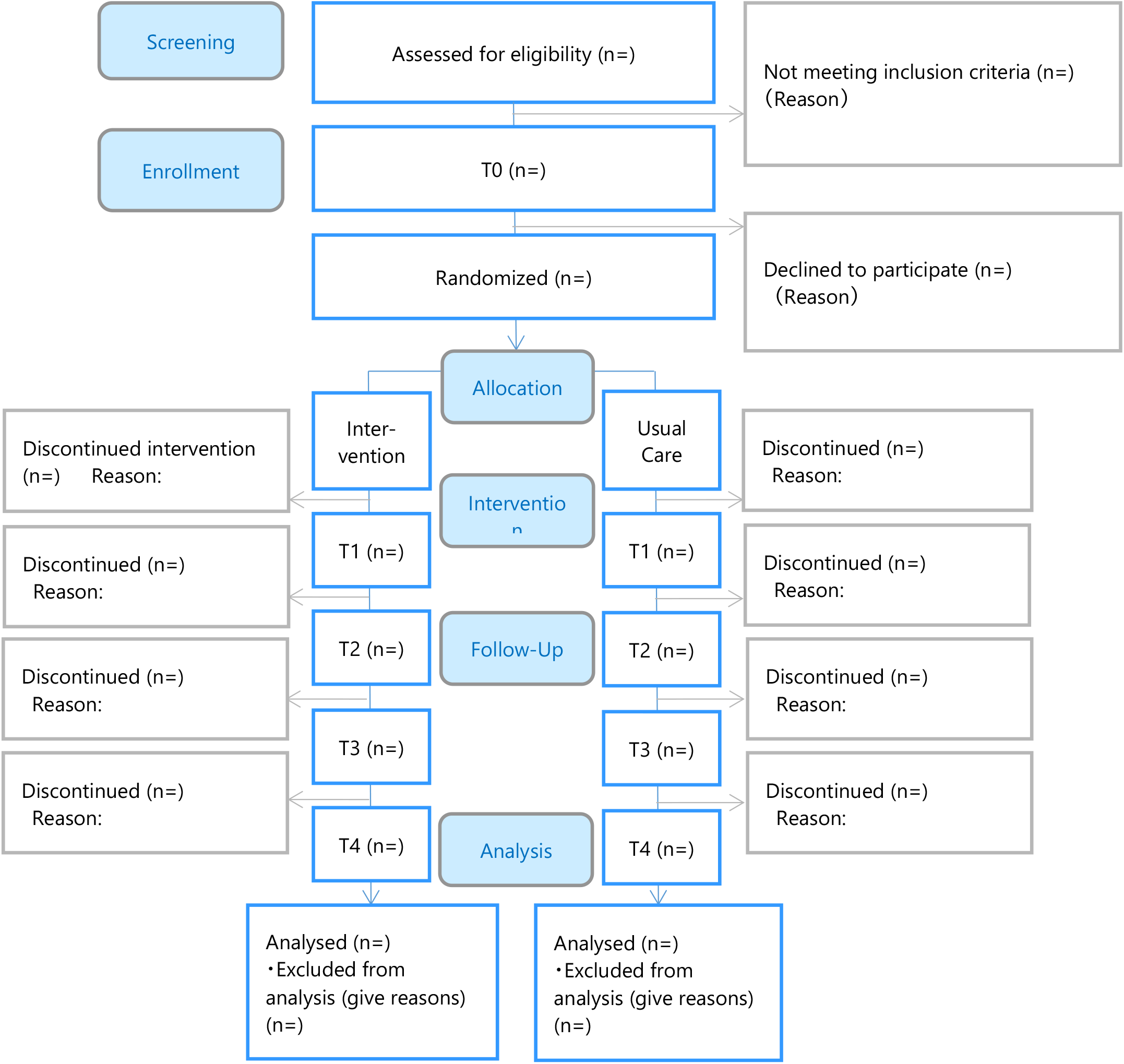
CONSORT diagram.

**Table 1.** Schedule for outcome measurement

|  | T0 | T1 | T2 | T3 | T4 |
| --- | --- | --- | --- | --- | --- |
| Outcomes | Baseline | Next oncologist visit scheduled after one week | Follow up at 1 week | Follow up at 12 weeks | Follow up at 24 weeks |
| **Primary outcome measure** |  |  |  |  |  |
| Oncologist's communication behaviors |  |  |  |  |  |
| SHARE score (RE subscale) |  | ○ |  |  |  |
| **Secondary outcome measures** |  |  |  |  |  |
| Oncologist's communication behaviors |  |  |  |  |  |
| SHARE score (S, H, and A subscales) |  | ○ |  |  |  |
| Communication behavior between participant and oncologist |  |  |  |  |  |
| Number of communication behaviors evaluated by RIAS |  | ○ |  |  |  |
| Number of conversations about ACP |  | ○ |  |  |  |
| Psychological distress |  |  |  |  |  |
| HADS | ○ | ○ | ○ | ○ | ○ |
| Quality of life |  |  |  |  |  |
| EORTC-QLQ-C30 | ○ |  | ○ | ○ | ○ |
| Participant care goals and preferred place for sending their final days |  |  |  |  |  |
| Care Goals and Preferred Place for Spending Their Final Days | ○ |  |  | ○ | ○ |
| Participant satisfaction with their oncologists' consultation |  |  |  |  |  |
| PSQ |  | ○ |  |  |  |
| Feasibility of the intervention |  |  |  |  |  |
| Usefulness, helpfulness, and comfort level of the intervention program |  | ◎ |  |  |  |
| Application log records |  |  |  |  | ◎ |
| **Demographics and clinical characteristics** |  |  |  |  |  |
| Medical care utilization |  |  |  |  | ○ |
| Medical and social background | ○ |  |  |  |  |

◎ Evaluated only in patients in the intervention group.
ACP, advance care planning; EORTC-QLQ-C30, European Organization for Research and Treatment of Cancer Quality of Life Questionnaire Core 30; HADS, Hospital Anxiety and Depression Scale; PSQ, Patient satisfaction questionnaire; RIAS, Roter interaction analysis system; SHARE communication model comprising four subscales categorized: S, Supportive environment; H, How to deliver bad news; A, Additional information; RE, Reassurance and Emotional support.

#### Intervention program

The intervention program, which is completed between T0 and T1, consists of two parts: QPL and identifying participants’ values (Table 2). Participants receive a brief explanation of the intervention program and how to use the app from an intervention provider. Participants are encouraged to complete all content on the app before an interview with an intervention provider. In the interview, an intervention provider reviews the items selected by a participant and assists them in considering priorities and verbalizing what is weighty to discuss with the oncologist. The interview is provided on the phone or face-to-face at the hospital, which is designed to take 30–60 minutes. In the outpatient visit following the interview, the intervention provider lets the oncologist know prior to the visit what the participant would like to discuss with their oncologist. Intervention providers are clinical psychologists, nurses, or psychiatrists who have participated in intensive training using the intervention manual. The intervention provider records and summarizes the intervention interviews, which are reviewed at weekly conferences.

**Table 2.** Intervention Program (QPL + Identifying patients’ value)

| Contents | Component Descriptions |
| --- | --- |
| QPL part with 45 questions categorized into the 8 topics | Eight topics (number of items for each topic):<br>1. Diagnosis and stage of disease (4)<br>2. Current treatment (7)<br>3. Symptom management and palliative care (4)<br>4. Future treatment (6)<br>5. Future living arrangements (9)<br>6. When standard treatment is no longer available (7)<br>7. Prognosis for the future (5)<br>8. Support for family (3). |
| Identifying the participant's values part | Three questions:<br>1. Things you value in terms of treatment and spending your days.<br>Question-1: This is a list of common examples of things people value in terms of treatment and spending the last days. Please select the one (or more) that you feel you would value.<br>Options: 18 domains of the Good Death Inventory (e.g., "Physical and psychological comfort" "Not Being a burden to others" "Good relationship with family")<br>2. Goals in terms of treatment and spending the last days developed based on the Goal Concordant Care framework.<br>Question-2: Please think about if you were to become ill or have difficulty continuing anticancer treatment as recommended by your doctor, then think about your goals for your further treatment and how you would like to spend your days. The following are some general examples of goals for treatment and spending time. Please choose one that most closely matches your idea.<br>Options: 1) I would like to receive treatment that relieves symptoms so that I can live a peaceful life, but I do not want to receive any cancer treatment that has any side effects or burdens, 2) I would like to receive cancer treatment that has few side effects and burdens so that I can continue my life as before, 3) I have important things I need to do, so I would like to receive cancer treatment even if there are side effects or burdens so that I can accomplish them, and 4) I would like to receive all cancer treatments, no matter what side effects or burdens they may cause, so that I can live as long as possible.<br>3. Places to spend the last days:<br>Question-3: choose where they would like to spend their days<br>Options: home, hospital near their home, palliative care unit/hospice, hospital they are visiting, or other. |

### Assessment measures

Table 1 shows the schedule for outcome measurement.

#### Primary outcome measure

##### The score of oncologists’ communication behaviors (reassurance and emotional support subscale from the SHARE scoring manual)

The conversation between the participants and the oncologists at visit T1 is audio-recorded, and the oncologist’s communication behavior is scored using the SHARE scoring manual (Table 3). SHARE is a conceptual communication skills model comprising 26 items and four subscales: S (s*upportive environment*; 2 items), H *(how to deliver bad news*; 7 items), A (*additional information*; (8 items), and RE (*reassurance and emotional support*; 9 items). We focus on RE, which assesses oncologists’ behavior in providing reassurance and their empathetic responses to participants’ emotions. [32] Scores range from 0 (*not applicable at all*) to 4 (*strongly applicable*). Scoring is conducted by multiple evaluators blinded to the assignment. Evaluators will be trained in conversation analysis with a manual, and inter-evaluators and intra-evaluators agreements will be checked in advance. We will adopt the items with a match rate ≥80%.

**Table 3.**
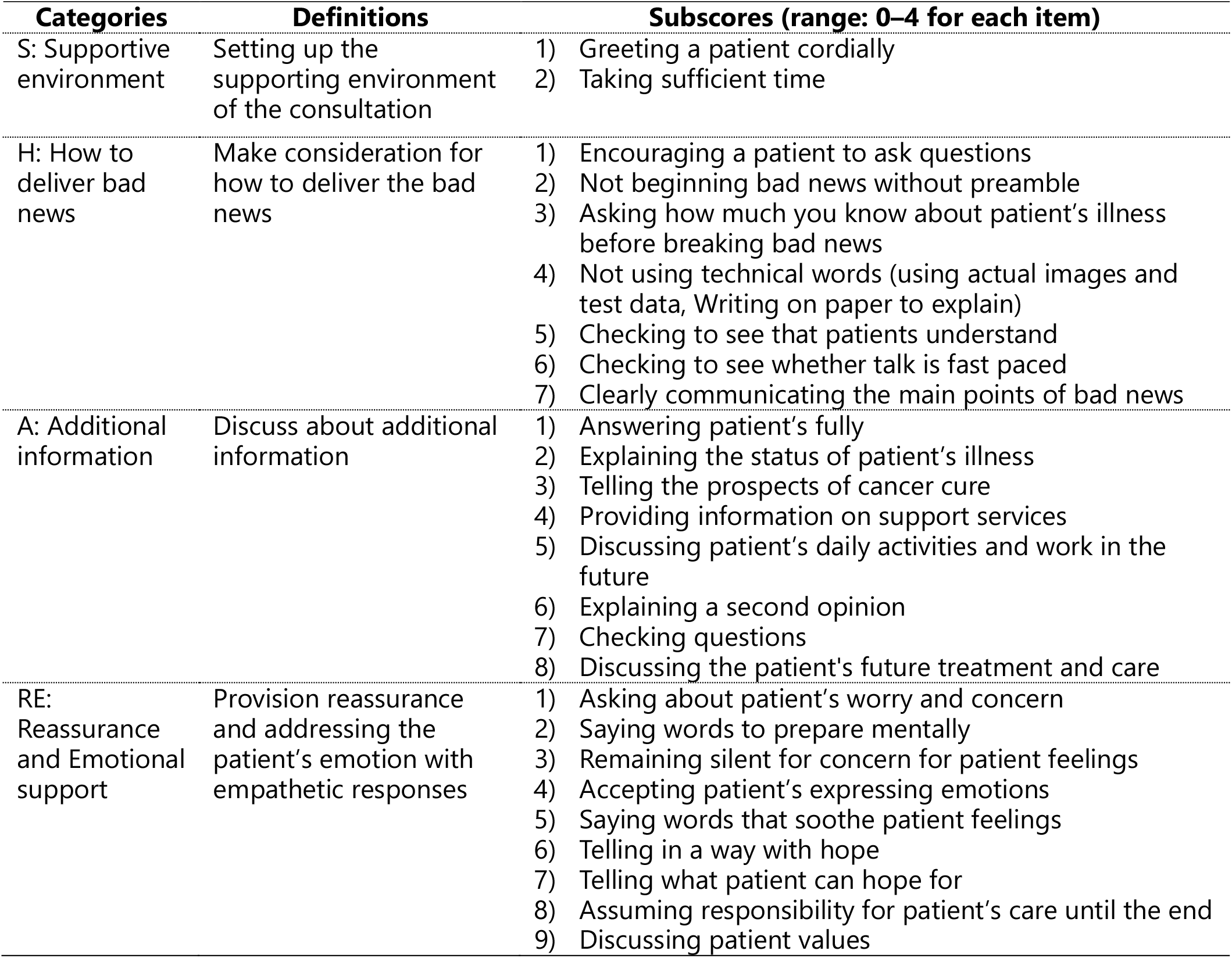
Oncologists’ communication behaviors: the SHARE coding manual

| Categories | Definitions | Subscores (range: 0–4 for each item) |
| --- | --- | --- |
| S: Supportive environment | Setting up the supporting environment of the consultation | 1) Greeting a patient cordially<br>2) Taking sufficient time |
| H: How to deliver bad news | Make consideration for how to deliver the bad news | 1) Encouraging a patient to ask questions<br>2) Not beginning bad news without preamble<br>3) Asking how much you know about patient's illness before breaking bad news<br>4) Not using technical words (using actual images and test data, Writing on paper to explain)<br>5) Checking to see that patients understand<br>6) Checking to see whether talk is fast paced<br>7) Clearly communicating the main points of bad news |
| A: Additional information | Discuss about additional information | 1) Answering patient's fully<br>2) Explaining the status of patient's illness<br>3) Telling the prospects of cancer cure<br>4) Providing information on support services<br>5) Discussing patient's daily activities and work in the future<br>6) Explaining a second opinion<br>7) Checking questions<br>8) Discussing the patient's future treatment and care |
| RE: Reassurance and Emotional support | Provision reassurance and addressing the patient's emotion with empathetic responses | 1) Asking about patient's worry and concern<br>2) Saying words to prepare mentally<br>3) Remaining silent for concern for patient feelings<br>4) Accepting patient's expressing emotions<br>5) Saying words that soothe patient feelings<br>6) Telling in a way with hope<br>7) Telling what patient can hope for<br>8) Assuming responsibility for patient's care until the end<br>9) Discussing patient values |

#### Secondary outcome measures

##### Oncologists’ communication behaviors

Oncologists’ communication behaviors at visit T1 are evaluated using the S, H, and A subscales of the SHARE manual. The scoring method is the same as for the RE subscale used in the primary outcome.

##### Communication behaviors between participants and oncologists

The audio-recorded conversations between the participant and oncologists are coded and the communication behaviors are counted using a computer version of the RIAS (the Roter interaction process analysis system). [33] The system is widely used in the US, the UK, and Japan. [34 35] Manuals have been translated into Japanese and validated for use in examining cancer patients. [36]

RIAS has 42 categories for coding the in-consultation communication behaviors. Two blinded trained coders assign one of the 42 codes to each utterance of the participants and the oncologists. To facilitate data interpretation, 21 categories related to the communication behaviors of interest in this study are grouped into four clusters based on the conceptual communication skills model used in previous studies. [32 37] Table 4 shows the categories that make up each of these clusters, and all RIAS categories are shown in the Appendix. The number of utterances in each cluster is also evaluated. Coders are trained and certified at the official training site, the RIAS Study Group Japan Chapter. Ten percent of the total consultations (25 consultations) will be double-coded and inter-coder reliability is examined regarding the degree of agreement for the identification of utterances and coding of each utterance. It should be verified in advance during the training period that the correlation coefficient meets 0.8. The reliability proved to be high (0.7–0.8) in previous studies. [34 38]

**Table 4.** Communication behaviors of both participants and oncologists: the Roter interaction process analysis system

| **RIAS clusters (N of categories)** | **Definitions** | **Categories** |
| --- | --- | --- |
| Setting up the interview (1) | Social behavior | Personal remarks and social conversation |
| Reassurance and empathic response (9) | Emotional responses, | Empathy<br>Legitimizing<br>Asks for reassurance<br>Showing partnership<br>Agreement<br>Encourages or shows optimism<br>Concern and worry<br>Approval<br>Asks psychosocial feelings |
| Medical and the other information giving (4) | Providing information related to medical care | Information giving:<br>Medical condition<br>Therapeutic regimen<br>Psychosocial feelings<br>Counseling (oncologist only):<br>Medical condition/therapeutic regimen |
| How to deliver the bad news (7) | Attitudes when communicating bad news | Question asking (open-ended):<br>Medical condition<br>Lifestyle information<br>Orientations and instruction<br>Asks for opinion<br>Asks for permission<br>Asks for understanding<br>Paraphrasing or checking |

RIAS, Roter interaction analysis system

##### Number of conversations about ACP

The number of conversations about ACP (prognosis, palliative care, remaining anticancer treatment, end-of-life treatment, recuperation issues, and preparation for the future) during the consultation is counted based on a conversation analysis manual developed by the previous study. [19]

##### Psychological Distress

Psychological distress is obtained at all five scheduled visits. The Hospital Anxiety and Depression Scale (HADS) is a 14-item self-report questionnaire developed for patients with medical illnesses. [39] It consists of anxiety and depression subscales (0 to 21 points each) with a 4-point scale, with higher scores indicating greater anxiety and depression. The Japanese version of the HADS has been validated in a cancer patient population. [40]

##### Quality of Life

Quality of life is obtained at T0, T2, T3, and T4. The European Organization for Research and Treatment of Cancer Quality of Life Questionnaire Core 30 (EORTC-QLQ-C30) is a four-domain, 30-item questionnaire consisting of functional scales, global health, and quality of life scales, symptom scales/items, and financial impact. [41] Scores for all scales range from 0 to 100. A high score on the functional scale would indicate high functioning, a high score on the global health and quality of life scale would indicate high health status, while a high score on the symptom scale and financial impact would indicate a high level of symptoms or problems. The reliability and validity of the Japanese version have already been confirmed. [42]

#### Participants’ care goals and preferred places for spending their final days

Participants will be asked about their goals and the places where they would prefer to spend their final days at T0, T3, and T4. We developed two original scales based on the conceptual diagram of care consistent with incurable cancer patient’s goals presented by Halpern [26] to assess 1) participants’ preferred treatment options after completion of standard care (care goal) and 2) participants’ preferred place where they would spend their final days. The treatment options are as follows: 1) I would like to receive treatment to alleviate my symptoms without any cancer treatment so that I can live a peaceful life, 2) I would like to receive less burdensome cancer treatment to alleviate symptoms so that I can continue my activity of daily living as much as possible, 3) I would like to receive cancer treatment even if it is somewhat burdensome so that I can finish crucial events or things that I have to do, 4) I would like to receive all cancer treatments, no matter what the burden, to prolong my life even for a day. The options for participants’ preferred place where they would spend their final days are as follows: 1) home, 2) a nearby hospital, 3) a palliative care hospital or ward, 4) the hospital where they are receiving treatment, and 5) others. The proportion of participants preferring unnecessarily aggressive treatment or impractical places of care is observed in each survey visit in an exploratory manner. We chose these outcomes for their comparability with previous studies. [5 43]

#### Participant satisfaction with their oncologists’ consultation

The Patient Satisfaction Survey (PSQ) [38 44 45] is conducted at T1. The 11-point scale (0, *not satisfied at all*, to 10, *very satisfied*) measures five categories of satisfaction with their oncology consultations: 1) needs addressed, 2) active involvement in the interaction, 3) adequacy of information, 4) emotional support received, and 5) the interaction overall. [43]

### Feasibility of the intervention

The timing of each data collection is shown in Table 1. The intervention’s feasibility is evaluated according to the participants’ assessment of the app’s usability, the time taken for interventions, and app log records. The app’s usability is determined by the following five questions: 1) Were the questions you wanted to ask identified by the time you saw your oncologist? 2) Did you understand and use the app? 3) Was the app program helpful? 4) Were you comfortable with the app program? 5) Was the telephone or in-person assistance helpful?

Participants rate each item on an 11-point scale (0, *not satisfied at all*, to 10, *very satisfied*). The intervention provider records the time taken for the intervention on the intervention report form. App log records, including time spent browsing and the operation status of the intervention program, are provided by the app developer.

### Medical care utilization

Medical care utilization is obtained from the electrical medical record of each participant at the 6-month follow-up. If the participant is not alive at 6 months, a medical record survey based on information at the time of death will be conducted. We will obtain the presence or absence of anticancer treatment and a reason for termination of treatment if the treatment is discontinued, unscheduled outpatient visits, hospitalization, ICU admission, use of end-of-life care consultations, and use of palliative care services.

### Medical and social background

The participant’s medical and social background information includes cancer type, length of time since diagnosis, age, gender, educational background, employment history, financial status, marital status, household status (lives with others, such as children or those requiring nursing care), methods and times of hospital visits, and whether there is a family member or other person who can accompany them.

### Harms

No particularly serious physical adverse events are anticipated for participants in this study. However, using the app may cause a psychological burden as participants think about preparing for when they will have difficulty continuing cancer treatments. Hence, newly diagnosed anxiety disorders or depression resulting from a psychological burden caused by the intervention will be considered an adverse event. If a participant reports that the intervention is causing a psychological burden or requests discontinuation of the intervention, the intervention will be stopped and the event will be reported promptly to their attending oncologists. Participants in the intervention group are scheduled to see an oncologist within one week after the intervention. Researchers will regularly check for updates to their medical records, if necessary, and case reports provided at regular team meetings so researchers can review the course of psychological distress, discuss changes in participants’ conditions caused by the intervention, and determine what should be reported to their attending oncologists.

### Compensation

Any unexpected health problems participants may experience due to their participation in this study will be adequately treated based on standard medical care covered by public health insurance programs, such as National Health Insurance. Participants receive a gift card worth 500 Japanese yen at T1.

### Sample size calculation

The results of a previous preliminary study showed that the effect size of the primary endpoint was 3.1. [22] In this study, the principal investigators agreed that an effect size of 2.5 would be considered clinically meaningful, given that this is an app-based intervention. Based on a significance level of two-sided 5% and a power of 80%, 250 subjects would be required. Since the National Cancer Center Hospital has many clinical trials, it may not be allow to participate in this study due to the conditions required of other clinical trials. If the participants overlap with a clinical trial, they may drop out of this study. At the National Cancer Center Hospital, the participation rate of patients who would be eligible for the study is approximately 30%. Therefore, the planned enrollment is 264 patients, assuming a 5% dropout rate due to clinical trial enrollment and other reasons during this study.

### Statistical Analysis

We will estimate the point estimates and 95% confidence intervals of the mean for each group and between-group differences for the primary endpoint. Two-tailed tests will determine significance at 5%. We will conduct the analysis using a general linear model with the clinical department, gender, and age as the adjustment factors for allocation. If the number of cases in each stratum is small, we will consider whether to adopt all adjustment factors. We will use a full analysis set consisting of the registered participant population who received at least part of the protocol treatment; however, participants deemed ineligible for the study after registration will be excluded from the analysis set. All statistical procedures, including the secondary endpoint and handling of missing data, will be detailed in the statistical analysis plan before data evaluation. The occurrence of discontinued cases after randomization will be assessed in both groups. Due to the nature of the intervention, the program may cause psychological burdens for some intervention group patients experiencing deteriorating physical conditions. Thus, patients’ reasons for discontinuation must be obtained (to the extent possible) so potential bias can be examined

### Data monitoring and management

An independent data monitoring team will report monitoring results semi-annually. The PRO data obtained will not be reported to individual participants or their oncologists to improve clinical care. Weekly meetings will be held between the research office and the monitoring team to discuss the progress of case enrollment and to report on cases. Data monitoring will be conducted using the entry data in EDC, Viedoc 4 (Viedoc Technologies, Sweden) and the central registration system by SUSMED, Inc (Tokyo, Japan). All paper data related to the study, including research assistant notes, intervention case reports, patient-reported questionnaires, and consent forms, will be stored securely in a lockable cabinet in the principal investigator’s office, as will audio-recorded data stored on an encrypted external hard drive. Only authorized researchers directly involved in the study will have access to the data. All data supporting the study results will be stored for a minimum of five years and will be available upon request to the corresponding author. A data monitoring plan is developed and kept by the data management team. No audit is required, and no data monitoring committee will be established. No interim analysis is planned.

## ETHICS AND DISSEMINATION

The study protocol was reviewed and approved by the Scientific Advisory Board of J-SUPPORT (registration No. 2104) and by the Institutional Review Board of the National Cancer Center Hospital (registration No. 2020-500). If significant protocol modifications are necessary, the investigators will discuss and report them to the committee for approval. The study will be conducted following the ethical guidelines for clinical studies published by the Japanese Ministry of Education, Science and Technology and the Ministry of Health, Labour and Welfare, the modified Act on the Protection of Personal Information, and the principles of the Declaration of Helsinki. The results of the RCT will be published in peer-reviewed scientific journals and presented at scientific meetings. After completing this RCT, our team will explore possibilities for expanding the app’s availability.

### Trial status

The study is currently recruiting participants; enrollment is scheduled through March 2023, with follow-up through September 2023.

## DISCUSSION

This study uses the mobile app to improve communication between patients and healthcare providers regarding ACP. Although the apps for behavior change and psychological intervention are increasing, this study is unique in its focus on facilitating communication related to ACP. Enhancing patients’ autonomous motivation is one of the mechanisms that can improve ACP discussion. [46] The advantage of the app program is that participants can find an environment and time where they can relax and actively engage in ACP. This is especially significant for cancer patients in the ACP program who need to think about their future treatment and life and express their values and what is crucial to them. The scoping review by McMahan et al. reported a lack of studies on healthcare systems and policies in the context of ACP. [5] It is expected that a healthcare system will be constructed so that ACP can reach the overall population in need. [47] The strength of ACP implemented with apps is the ease of adaptation to the healthcare system, which is promising in a world where COVID-19 situations are uncertain.

For the evaluation of ACP discussions, there is currently no gold standard for assessing discussions between patients and healthcare providers and their outcomes. Previous studies have used family assessments [15 20] and the originally developed composite communication assessment. [19] We expect the outcomes of this study will provide multiple communication components confirming the facilitation of various aspects of ACP discussions. We hypothesized that introducing ACP discussions early in cancer treatment would improve communication with healthcare providers and lead to ongoing discussions, which would help improve health outcomes and end-of-life care. However, these outcomes could only be evaluated in an exploratory manner in this study, and it is urgent to evaluate patient-centered outcomes, such as goal-concordant care, in the future. [43]

Although the eligibility criteria are based on ACP guidelines, depending on the participant’s readiness, some participants may feel it is too early to consider future treatment and end-of-life while undergoing cancer treatment. There has been much discussion about the appropriate timing of ACP, which is likely to be triggered by a patient’s deteriorating health or reduced treatment options. [48] However, there is no evidence regarding the appropriate timing for introducing ACP discussions, [48] and it is assumed that some participants may find this intervention burdensome. Moreover, healthcare providers might hesitate to initiate the discussion for fear of causing anxiety to the patient; thus, more careful ACP referrals and a qualitative exploration of study dropouts are required.

The study has several methodological limitations. Although not all eligible patients may own a mobile device compatible with the app, we determined device access would not limit eligibility. Hence, to allow for a diverse group of participants, iPads able to run the program app were on loan as alternative means of participation. While patients unfamiliar with app use could participate in this study, consideration should be given to patients who are unable to use the app when adapting to the real world.

Second, the intervention package consists of multiple components, including the introductory session with the app, and patients’ choice of questions to ask and share with their oncologists. We cannot indicate which components are most effective in improving communication. Individualized evaluation of app usage, intervention adherence, and patient satisfaction should be conducted to understand the challenges ahead for the next step.

Finally, we hypothesize that the intervention program will improve communication between patients and oncologists, leading to ongoing discussions and improving the quality of end-of-life care, but with the limitation that it is a partial and indirect evaluation of ACP. Although the primary outcome was selected after careful consideration, there is no established method for evaluating ACP, and standardized measurement is still a challenge.

## Data Availability

The study protocol, data definition tables, and dataset will be uploaded to the UMIN-Individual Case Data Repository

https://www.umin.ac.jp/icdr/index-j.html.

## Acknowledgments

We thank Ms. A. Akama and Mr. N. Akama for their support with data management. We thank Dr. K. Sudo, Dr. C. Morizane, Dr. T. Yoshida, Ms. T. Mashiko, Mr. Y. Watanabe, Mr. T. Motohashi, and Dr. Shimazu for their advice about our study. We also thank Ms. K. Shinozaki, Ms. M. Okubo, Ms. I. Tanaka, Ms. H. Kouno, Ms. H. Masubuchi, Ms. C. Kida, Ms. K. Hata, Ms. A. Sato, Mr. S. Goto, and Ms. M. Kanamaru for data collection and logistic assistance.

## Authors’ contributions

YU is the principal investigator. YU and MF conceived the concept of the study. YU, MF, NB, TA, MM, NS, TU, AO, TM, and MK were involved in the study design. TY and SO developed the statistical analysis plan. AO and TM played roles in data management. KO, MK, MO, and MF contributed to data collection. YU, MF, and MO supervised the conduct of this study. KO, MO, and MF drafted the manuscript. YU, NB, TA, MM, and the other authors contributed substantially to the revision of the manuscript. All authors have approved the manuscript as submitted and agreed to accept responsibility for any part of the work.

## Competing interests

Competing interests: All authors declare that they have no competing interests regarding this work. BN reports grants from Ono Pharmaceutical and Takeda Pharmaceutical, and personal fees from Ono Pharmaceutical, Bristol-Myers Squibb, Daiichi Sankyo, and Taiho Pharmaceutical. TA reports grants from Daiichi Sankyo, Eisai, Fujifilm RI Pharma, MSD, Otsuka Pharmaceutical, and Shionogi, personal fees from Igaku-Shoin, AstraZeneca, Chugai Pharmaceutical, Daiichi Sankyo, Sumitomo Dainippon Pharma, Eisai, Janssen Pharmaceutical, Kyowa Kirin, Eli Lilly, MSD, Meiji Seika Pharma, Mochida Pharmaceutical, NIPRO, Nippon Zoki Pharmaceutical, Otsuka Pharmaceutical, Pfizer, Takeda Pharmaceutical and Viatris, and pending patents (2019-017498 & 2020-135195). TY reports grants or contracts from AC Medical Inc., A2 Healthcare Corporation, EP Croit Co., Ltd., ClinChoice., Japan Tobacco Inc., Japan Media Corporation, Medidata Solutions, Inc., Ono Pharmaceutical Co., Ltd., Asahi Intecc Co., Ltd., 3H Clinical Trial Inc., Medrio, Inc., Nipro Corporation, Intellim Corporation, Welby Inc., 3H Medi Solution Inc., Nipro Corporation, BaseConnect Inc., Nobori Ltd., Puravida Technologies LLC., and Hemp Kitchen Inc. and grants to the affiliated institutions from Kyowa Kirin Co., Ltd., Tsumura & CO., Daiichi Sankyo Company, Limited., Otsuka Pharmaceutical Co., Ltd., and Eisai Co., Ltd., and consulting fees from EPS Corporation., Japan Tobacco Inc., Medidata Solutions, Inc., Ono Pharmaceutical Co., Ltd., Kowa Company, Ltd., Chugai Pharmaceutical Co., Ltd., Tsumura & Co., Daiichi Sankyo Company, Limited., Eisai Co., Ltd., Asahi Intecc Co., Ltd., Asahi Kasei Pharma Corporation, 3H Clinical Trial Inc., Intellim Corporation, Takeda, AstraZeneca, Sonire Therapeutics Inc., Seikagaku Corporation, and Merck & Co., Inc., and personal fees from Nipro Corporation.

### Trial registration number

UMIN000045305; NCT05045040

---

**Table A1.**
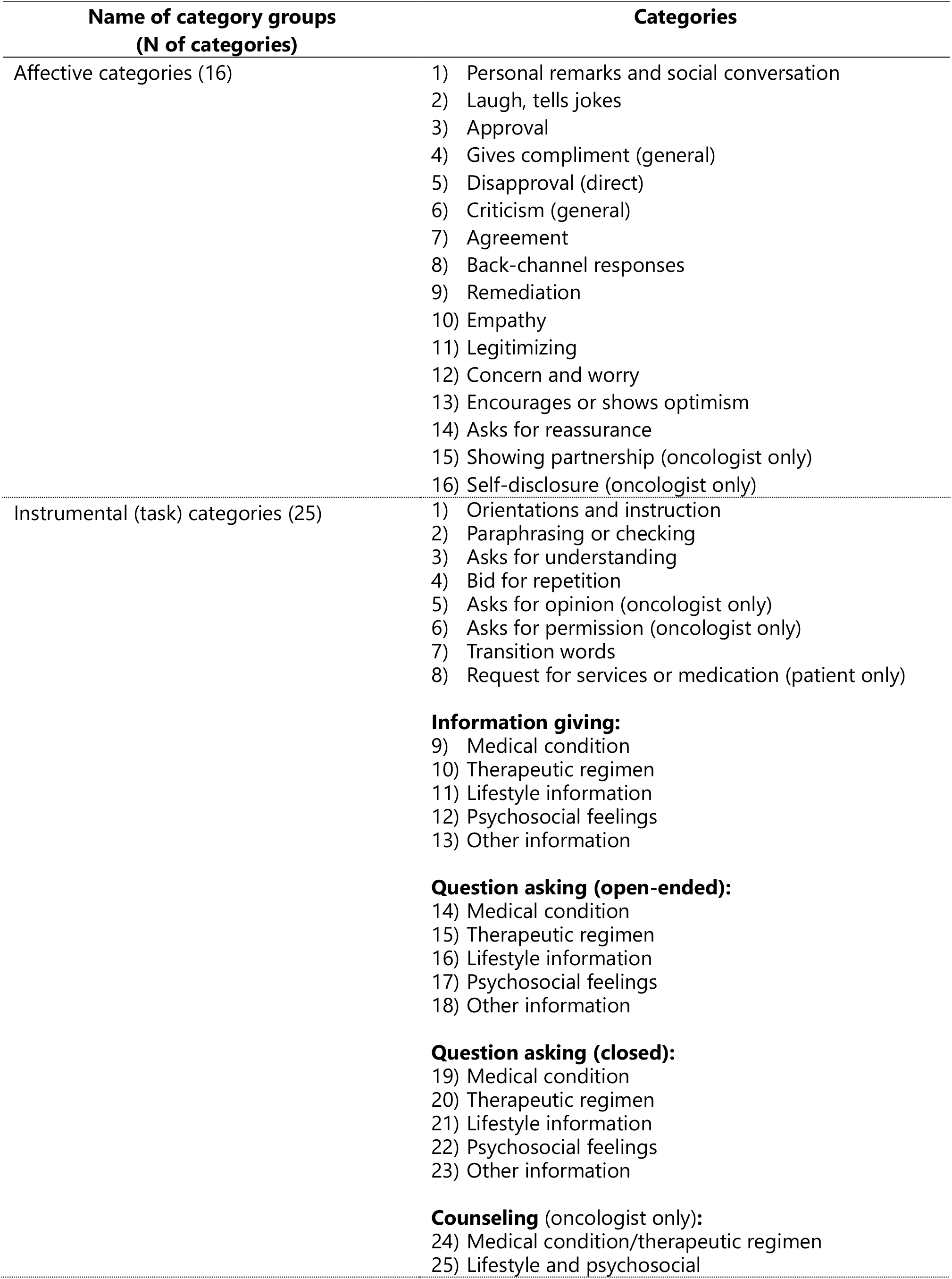
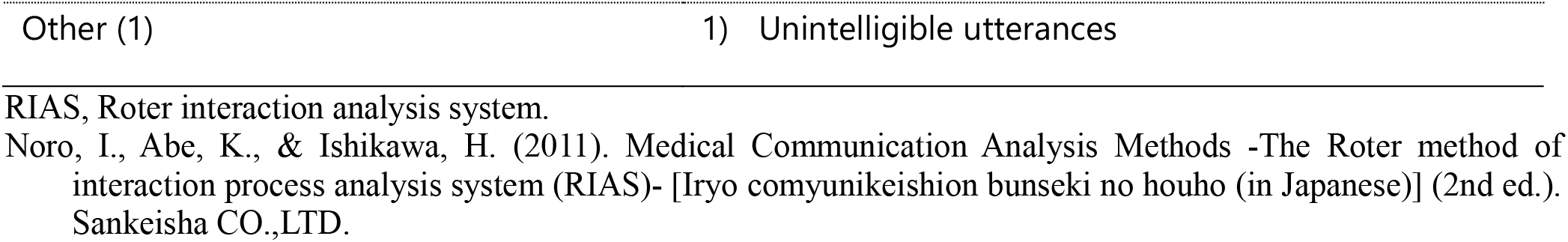
The RIAS all categories (Noro et al., 2011)

